# Exploring COVID-19 Pandemic Perceptions and Vaccine Uptake among Community Members and Primary Healthcare Workers in Nigeria: A Mixed Methods Study

**DOI:** 10.1101/2024.09.02.24312966

**Authors:** Abiodun Sogbesan, Ayobami Bakare, Sibylle Herzig van Wees, Julius Salako, Damola Bakare, Omotayo E Olojede, Kofoworola Akinsola, Oluwabunmi R. Bakare, Adegoke Falade, Carina King

## Abstract

**Background:** The COVID-19 pandemic significantly impacted global health, with diverse perceptions about the disease and control measures, including vaccination. Understanding these perceptions can help inform public health and vaccination strategies in future outbreaks. This study examined community members and healthcare workers’ (HCWs) perceptions of the COVID-19 pandemic and vaccines in Nigeria, exploring factors influencing vaccine acceptance and hesitancy.

**Methods:** We conducted a mixed-methods study, combining quantitative survey data from 2,602 respondents (2,206 community members and 396 HCWs) with qualitative interviews. Quantitative data were analyzed to identify factors associated with COVID-19 vaccine uptake and pandemic perceptions, while qualitative insights provided a deeper understanding of cultural perceptions, experiences, and hesitancy towards the COVID-19 vaccine.

**Results:** Overall, 43.4% of community members and 96.7% of HCWs received at least one dose of COVID-19 vaccine. Vaccine uptake was positively associated with increasing age, previous COVID-19 testing, male sex, government employment, and knowing someone diagnosed with COVID-19. Christianity was associated with lower uptake among community members. Perceptions varied, with 34.2% of community members and 17.7% of HCWs considering COVID-19 a death sentence, while 27.8% and 22.0% believed in ‘African immunity,’ respectively. Hesitancy was driven by the fear of side effects (32.6%), pregnancy-related concerns (25.9%), and convenience-related issues (13.5%). Qualitative data found misinformation, mistrust, fear of adverse reactions, logistical challenges, and belief in the sufficiency of childhood vaccination fuelled hesitancy towards the COVID-19 vaccine. Despite this, general trust in vaccine safety and efficacy remained high, with most respondents willing to be vaccinated against other diseases and future outbreaks.

**Conclusion:** Our findings underscore the need for tailored public health strategies to address specific sociodemographic factors, individual perceptions, and logistical challenges to enhance COVID-19 vaccine uptake. Public health campaigns should focus on debunking myths, improving vaccine literacy, and leveraging the social influence of respected community leaders to build trust.

## Introduction

Vaccines have contributed tremendously to improving global health, and their implementation is one of the most reliable and cost-effective interventions in public health that continue to save millions of lives every year. (1–3) However, vaccine hesitancy is one of the top 10 global health threats(4), and was prominent during the COVID-19 pandemic. After the discovery of the genome sequence of SARS-CoV-2 in early 2020, many COVID-19 vaccines were developed, with 50 of these vaccines receiving approval in 201 countries and 11 listed for emergency use by WHO.(5–9) Subsequently, a global COVID-19 vaccination target was set for countries to vaccinate at least 70% of their populations by mid-2022.(9) Vaccination of 100% of health workers and 100% of the most vulnerable groups, with people aged over 60 years old and those who are immunocompromised or have underlying health conditions, is expected to be prioritized.(9)

Despite COVID-19 vaccine roll-out and administration across all regions of the world, only 66.1% of the eligible population had been vaccinated with a complete primary series as of 30 August 2023.(10) While the Western Pacific (85.4%) and the Americas (71.2%) have surpassed the 70% vaccination target, the WHO African region has lagged behind, with only 32.4% fully vaccinated.(10) Therefore, Africa has failed to achieve the African Union’s goal of 70% population vaccine coverage by the end of 2022.(7,10,11) In Nigeria, despite having seven government-approved vaccines, only 37.8% of eligible individuals have been fully vaccinated as of August 30, 2023.(10) This represents a shortfall in meeting both international and the Federal Government vaccination coverage goals of 40% and 70% by the end of 2021 and 2022, respectively.(12)

The reasons for low COVID-19 vaccination uptake are complex. The 3C model (convenience, complacency, and confidence) has been used to describe factors influencing COVID-19 vaccine uptake in sub-Saharan Africa.(13,14) Specifically, in Nigeria and across Africa, factors such as fears of side effects, ineffective public health communication, rumours and misinformation, and anxiety have contributed to low confidence in the safety and efficacy of COVID-19 vaccines.(15–17) Conversely, weaknesses in health systems, logistical gaps, inadequate funding, shortages of trained vaccinators, disruptions to essential health services, and concerns about vaccine accessibility impede the convenience and accessibility of COVID-19 vaccination efforts.(11,15,17,18) Finally, insufficient planning, apathy and disbelief in the existence of COVID-19 contributed to complacency or delays in COVID-19 vaccine administration.(11,15,17)

In Nigeria, the factors influencing vaccine decision-making have not been sufficiently researched, particularly in the context of the COVID-19 pandemic. There is also limited evidence regarding the perceptions of COVID-19 vaccines among communities and healthcare workers. Investigating healthcare worker perceptions is essential, as they may significantly influence the effectiveness of vaccination promotion within the community. Therefore, we aimed to examine vaccination patterns, barriers, and drivers of COVID-19 vaccine uptake among Nigerians. This study allowed us to explore the complexities inherent in COVID-19 immunization programmes, and to learn lessons that will be relevant in the context of a new pandemic.

## Methods

### Study design

We conducted a mixed-method parallel convergent study (QUAN + qual), comprising a facility-based cross-sectional quantitative survey and qualitative discussions with healthcare providers and community members. The study was carried out in three states in Nigeria, Oyo and Lagos in Southwestern Nigeria and Jigawa in Northwestern Nigeria from June 27 – September 13, 2022. This study is a component of a larger mixed-methods investigation, the ‘COVID-19 Vaccine Programme Delivery in Nigeria,’ which examined the perspectives of healthcare providers and community members regarding COVID-19 vaccination programme delivery, and how it exerts influence on and differs from the routine immunization programme, in Nigeria. Quantitative and qualitative results are presented together and triangulated under common headings.

### Study settings

The choice of Lagos, Oyo and Jigawa states was based on the COVID-19 burden and performance in routine and COVID-19 immunization programmes. Lagos and Oyo ranked among the top five states with the highest COVID-19 burden,(19) but were not among the top five performers during early phases of the COVID-19 vaccine rollout.(20) Jigawa, despite not having a high COVID-19 burden, was the second top-performing state for initial COVID-19 vaccine roll out.(20)

Oyo and Lagos states are predominantly inhabited by the Yoruba ethnic group, with Christianity and Islam as the dominant religions. Jigawa state is located in the Northwest geopolitical zone and predominantly inhabited by Hausa and Fulani ethnic groups, with Islam being the most practiced religion. Although Lagos is the smallest of the three states in terms of land mass, it has the highest population, with an estimated population of 24.6 million.(21) Oyo and Jigawa, on the other hand, have estimated populations of 7.84 million and 7.49 million, respectively.(22,23) The study was conducted in Egbeda and Ibadan Southwest Local Government Area (LGA) in Oyo state, Kiyawa and Dutse LGAs in Jigawa state, and Ikeja and Ikorodu LGAs in Lagos state. The LGAs were purposively selected based on their feasibility and accessibility.

### Study population

Data were collected from healthcare providers involved in immunization in the selected study sites. Community members included mothers who brought their children for routine immunization and adult recipients of COVID-19 vaccines in primary health facilities of the selected LGAs. Community members who recently relocated, were not LGA residents, or required urgent medical attention were excluded.

### Quantitative data collection and analysis

#### Sample size determination

The sample size for this study was determined based on the minimum sample size calculated in the wider study protocol for the Covid-19 Vaccine Programme delivery in Nigeria, based on estimating a single proportion: n = Zα^2^ (p*(1-p)/d^2^. P represents the proportion of healthcare workers reporting post-vaccination side effects in Enugu South-East Nigeria (p = 87.6%),(24) with a precision of 5% (d=0.05) and a confidence interval of 95% (z = 1.96). The estimated study sample was 558 for the three states after adjusting for 10% non-response.

#### Sampling technique

In each purposively selected LGA, we identified public and private health facilities that offered COVID-19 vaccination services and routine immunization from the federal government health facilities database. Facilities offering COVID-19 vaccinations, where possible, were matched in terms of geography and ownership with facilities that offered only routine immunization and/or outpatient services during the data collection period. In Lagos and Oyo, 11 PHCs offering COVID-19 vaccination and 11 PHCs offering routine immunization/outpatient services were randomly selected. In Jigawa, 11 primary healthcare facilities and 11 health posts offering COVID-19 vaccination were randomly selected for each LGA (n=88 facilities overall). All mothers and general adult outpatient participants who presented at the selected facilities were approached to participate using convenience sampling. In addition, all healthcare workers involved in immunization services in the selected facilities were purposefully selected.

#### Data collection

Data was collected using an interviewer-assisted questionnaire pre-tested in all three states. Trained data collectors with at least secondary education conducted in-person interviews to obtain information on respondents’ sociodemographic characteristics, perceptions about COVID-19, COVID-19 experiences, including uptake of vaccination and side effects, perception about COVID-19 vaccines, and willingness to take other vaccines. Sociodemographic information assessed were age, sex, religion, ethnicity, marital status, household wealth index, monthly income, employment status, and education. Data was collected on Android tablets using Open Data Kit (ODK) software, and regular checks were performed for accuracy.

#### Study variables

The primary outcome variable, “COVID-19 vaccine uptake,” was defined as self-reported receipt of any dose of COVID-19 vaccine. Exposures of interest were categorized as: level of education (no formal education, primary, secondary, and tertiary), religion (Christianity, Islam), and government employment (yes/no). Household wealth index was analyzed using principal component analysis and categorized into tertiles.

#### Data Management and Analysis

We performed all quantitative data analyses using Stata 16.0. We described respondents’ characteristics, perception of COVID-19 disease, COVID-19 experiences, including the pattern of vaccination, and reasons for COVID-19 vaccine acceptance and hesitancy, and perception about COVID-19 vaccine and willingness to take other vaccines using frequencies, percentages, means and standard deviation. We used multivariable logistic regression to assess respondent factors associated with COVID-19 vaccine acceptance.

### Qualitative data collection and analysis

#### Sample size and sampling

Purposive and convenience sampling techniques were employed to recruit 14 healthcare providers who were involved in the national vaccination program (Jigawa, 8; Oyo, 6) for in-depth semi-structured interviews. For the community members, we interviewed 16 individuals (Jigawa:8, and Oyo:8), with maximum variation sampling to include those who received zero, 1, and 2 doses. This sample size was determined based on the expectation that the number would be sufficient to achieve saturation after conducting 9-17 interviews.(25)

#### Interview guide

The interview guides were developed based on our literature review.(26,27) The interview guide for healthcare providers included four sections focused on the participants’ socio-demographic information, their perception of COVID-19 vaccination, their understanding of COVID-19 vaccines and vaccination doses, and their experience with COVID-19 vaccination (S5 Interview guide). The interview guide for community members had three sections focused on participants’ socio-demographic information, perception of COVID-19 vaccination, and vaccination experiences (S6 Interview guide).

#### Data collection

The research team comprised public health specialists. Interviews in Jigawa were conducted in English and Hausa by JS and two other female research nurses who have prior experience in qualitative data collection and are acquainted with the context. Interviews in Oyo were conducted in English and Yoruba by KOA and a research nurse with knowledge of the local setting. KOA is a female public health researcher with a Master of Public Health degree from Nigeria and experience in qualitative research. Interviews were conducted face-to-face in private locations for the participants and interviewers, which were home visits or workplaces for community members and primary healthcare facilities for healthcare workers. Each participant was given detergent as an incentive at the end of the interview. The interviews lasted between 45-60 minutes. Field notes were made during the interviews, and no repeat interviews were conducted.

#### Data management and analysis

Interviews were audio-recorded, transcribed, and translated verbatim in English and then stored in a secure cloud platform with restricted access to non-research team members. Thematic analysis was used to identify separate codes and themes for health providers and community members.(28) AAB and KOA coded the data double-blinded. After the first round of coding, the codebook was compared and discussed. The coding team initially categorized and developed themes derived from the data, which was then expanded to the entire research team.

#### Reflexivity

The interviewers were non-indigenes who had no prior relationship with the participants but spoke the same language as the participants. AAB and KOA demonstrated cultural sensitivity, recognizing that being based in Oyo state and understanding cultural nuances, societal norms, and beliefs could shape their interpretation of participants’ responses. They also acknowledged the potential influence of their professional roles, training, and experiences, including any preconceived notions about vaccination, on their understanding of their experiences. It was noted that AAB, as a male community health physician, might bring a clinical perspective to the analysis, emphasizing individual health behaviours, patient-provider interaction, and the impact of medical misinformation on vaccine acceptance. KOA, on the other hand, being a female public health researcher, approached the analysis from a broader public health perspective, focusing on systemic issues, including healthcare and socio-economic determinants, as well as public health interventions in promoting vaccine uptake. Continuous reflections on different backgrounds were discussed in a small team and the larger research team.

### Ethical consideration

Ethical approval was obtained from the relevant ethics committees in all three states, including the UI/UCH Ethics Committee (ref: UI/EC/22/0139), Oyo State Ministry of Health (ref: AD/13/479/44396A), Lagos State Government (LREC/06/10/1870) and Jigawa State Ministry of Health (ref: JGHREC/2022/093). The study was conducted in compliance with the Declaration of Helsinki and Nigerian National Code of Health Research Ethics. Verbal consent was obtained prior to the respondents’ participation in the quantitative study, while written informed consent was obtained for the qualitative study. They were given the opportunity to review the informed consent form. In both studies, participants were notified that their involvement was voluntary, and that the data collected would be used solely for research purposes.

## Results

### Study Participants

We included 2602 participants in the quantitative analysis, 2206 (84.8%) community members and 396 (15.2%) healthcare workers (S1 Fig.). The majority of the community members (83.9%) were female, with a mean age of 33.2 years (SD±11.2). Over half of the community members (55.0%) belonged to the Yoruba ethnic group, 55.0% practised Islam, 34.8% had secondary education, and 42.5% belonged to a ‘poor’ household. For healthcare workers, 81.1% were female, with a mean age of 36.7 years (SD±10.3). The majority of healthcare workers (72.0%) belonged to the Yoruba ethnic group, 58.3% practised Christianity, 83.3% had tertiary education, 50.5% belonged to ‘wealthy’ households, and 44.2% were not employed by the government despite working in public primary health facilities. (Table 1).

**Table 1:** Socio-demographic characteristics of respondents (N = 2602)

| **Characteristics** | **Community members/patients (N= 2206)** | **Healthcare workers (N=396)** |
| --- | --- | --- |
|  | **N (%)** | **N (%)** |
| **Age at recruitment (years)** |  |  |
| Mean (SD) | 33.2 (11.5) | 36.7 (10.3) |
| Minimum – maximum | 15 – 91 | 18 – 59 |
| **Sex** |  |  |
| Male | 354 (16.1) | 75 (18.9) |
| Female | 1852 (83.9) | 321 (81.1) |
| **Highest level of education** |  |  |
| No formal education | 474 (21.5) | 1 (0.2) |
| Primary | 239 (10.8) | 5 (1.3) |
| Secondary | 767 (34.8) | 58 (14.7) |
| Tertiary | 726 (32.9) | 330 (83.3) |
| Missing | - | 2 (0.5) |
| **Religion** |  |  |
| Christianity | 988 (44.8) | 231 (58.3) |
| Islam | 1213 (55.0) | 165 (41.7) |
| Other <sup>a</sup> | 4 (0.1) | - |
| Missing | 1 (0.1) |  |
| **Ethnicity** |  |  |
| Yoruba | 1213 (55.0) | 285 (72.0) |
| Hausa | 597 (27.1) | 72 (18.2) |
| Igbo | 163 (7.4) | 11 (2.8) |
| Fulani | 111 (5.0) | 10 (2.5) |
| Other | 122 (5.5) | 18 (4.5) |
| **Employment by government** |  |  |
| No | 2090 (94.7) | 175 (44.2) |
| Yes | 116 (5.3) | 221 (55.8) |
| **Monthly income (in naira)** |  |  |
| < 30,000 (< minimum wage) | 677 (30.7) | 62 (15.7) |
| ≥ 30,000 | 318 (14.4) | 109 (27.5) |
| Declined | 1211 (54.9) | 225 (56.8) |
| **Wealth index (in tertiles)** |  |  |
| Lowest | 937 (42.5) | 79 (19.9) |
| Middle | 468 (21.2) | 110 (27.8) |
| Highest | 733 (33.2) | 200 (50.5) |
| Missing | 68 (3.1) | 7 (1.8) |

<sup>a</sup>Three respondents practise traditional religion, and one respondent with no religion

For the qualitative interviews, we recruited more females than males across community members (13 of 16 participants) and healthcare providers (8 of 14 participants). All 6 male healthcare providers were recruited from Jigawa State (S2 Table).

### Socio-demographic factors influencing vaccine acceptance

Analysis of vaccine uptake patterns showed a clear difference between community and HCW uptake, with 43.4% (957/2206) of community members receiving at least one dose of the COVID-19 vaccine, compared to 96.7% (383/396) of healthcare workers (Table 2). The proportion of individuals who had previously been tested for COVID-19 was considerably lower in both groups, with only 9.3% (206/2206) of community members and 33.0% (132/396) of healthcare workers having undergone testing. Additionally, 14.2% (314/2206) of the community members and 29.0% (115/396) of the healthcare workers knew someone previously diagnosed with COVID-19 infection (Table 2).

**Table 2:**
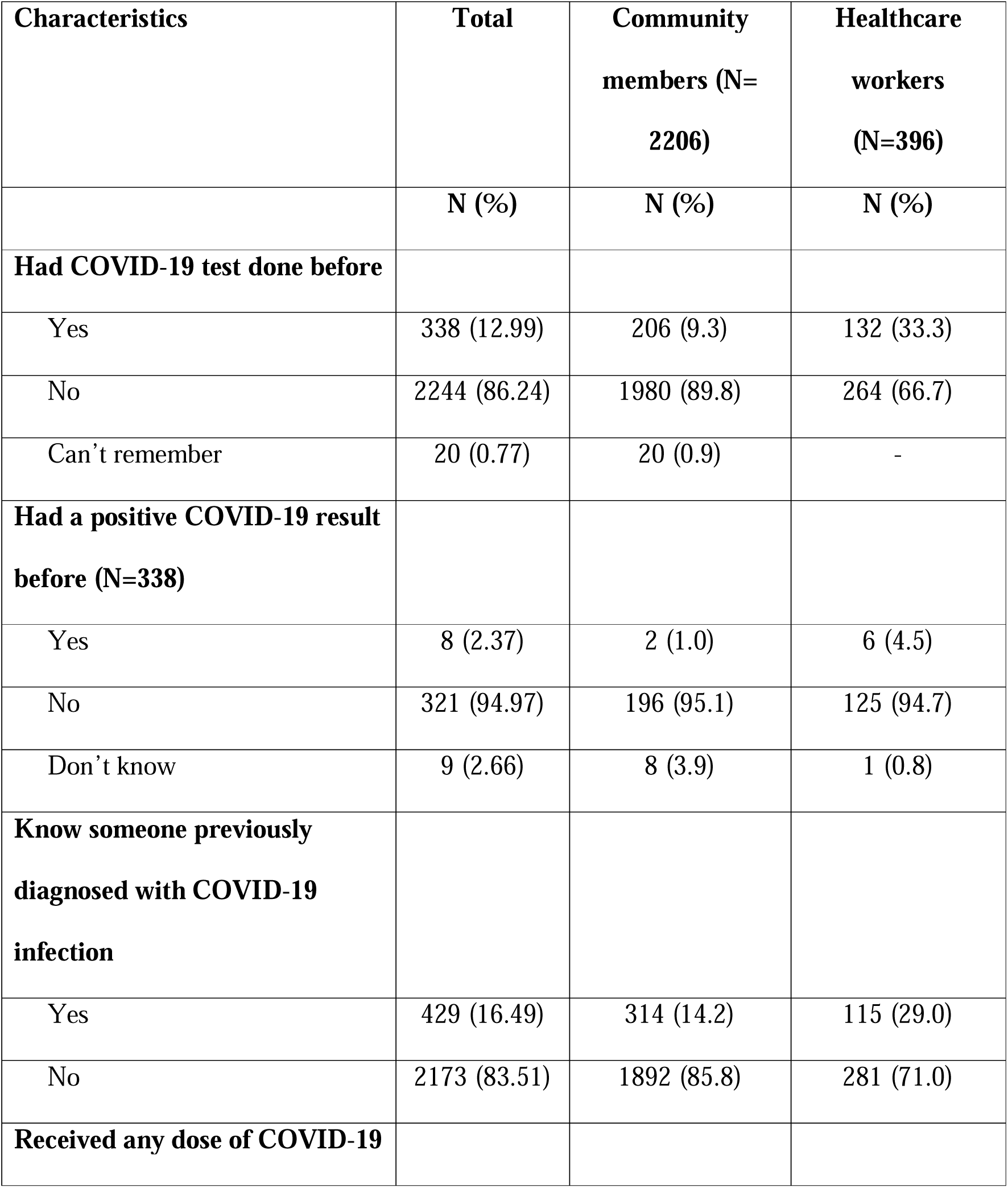
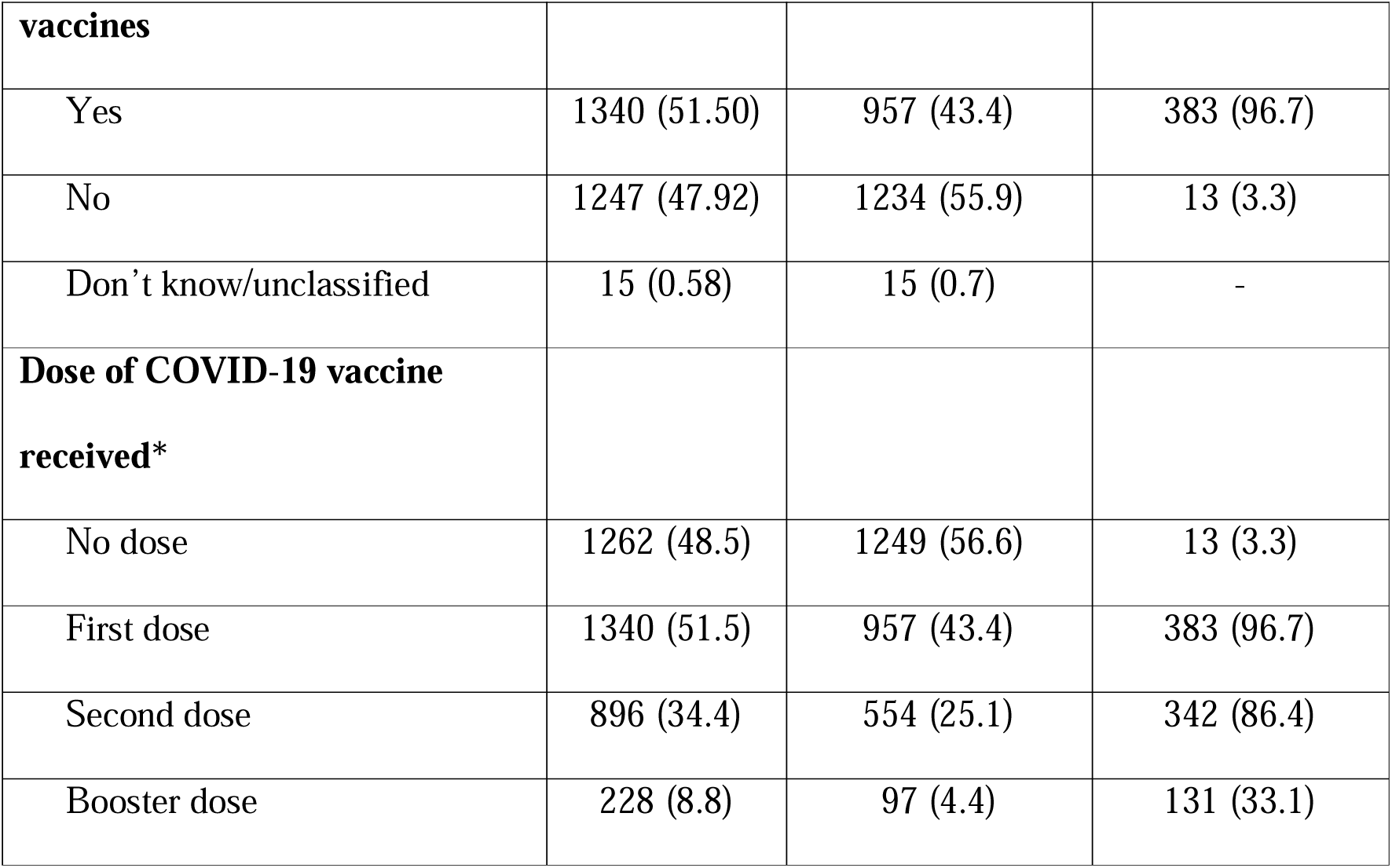
Respondents’ COVID-19 experience (N = 2602)

The odds of vaccination among community members and healthcare workers increased with increasing age [AOR = 1.04; 95% CI: 1.03 – 1.06] and [AOR = 1.06; 95% CI: 1.00 – 1.13], respectively, and previous COVID-19 testing [AOR = 4.32; 95% CI: 3.13 – 5.98], and [AOR = 8.25; 95% CI: 1.04 – 65.28] (Table3 and 4).

For community members only, being male [AOR = 2.68; 95% CI: 1.54 – 4.66], having government employment [AOR = 2.46; 95% CI: 1.34 – 4.52], and knowing someone diagnosed with COVID-19 [AOR = 1.41; 95% CI: 1.06 – 1.87] were also associated with higher odds of vaccination. Uptake was lower among community members who practised Christianity [AOR = 0.73; 95% CI: 0.55 – 0.97] (Table 3).

**Table 3:** Factors associated with COVID-19 vaccine uptake among community members.

| Characteristics | Crude OR (95% | p-value | Adjusted OR <sup>a</sup> | p-value |
| --- | --- | --- | --- | --- |

|  | **CI)** |  | **(95% CI)** |  |
| --- | --- | --- | --- | --- |
| **Age** | 1.05 (1.04 – 1.06) | **< 0.001** | 1.04 (1.03 – 1.06) | **< 0.001** |
| **Sex** |  |  |  |  |
| Female | 1 |  | 1 |  |
| Male | 3.96 (3.09 – 5.07) | **< 0.001** | 2.68 (1.54 – 4.66) | **< 0.001** |
| **Education** |  |  |  |  |
| Primary or less | 1 |  | 1 |  |
| Secondary | 1.25 (1.02 – 1.55) | **0.035** | 1.34 (0.69 – 2.60) | 0.387 |
| Tertiary | 1.95 (1.58 – 2.40) | **< 0.001** | 1.56 (0.82 – 2.96) | 0.172 |
| **Employed by government** |  |  |  |  |
| No | 1 |  | 1 |  |
| Yes | 5.14 (3.28 – 8.08) | **< 0.001** | 2.46 (1.34 – 4.52) | **0.004** |
| **Wealth index** |  |  |  |  |
| **Lowest** | 1 |  | 1 |  |
| Middle | 1.27 (1.02 – 1.60) | **0.036** | 0.88 (0.71 – 1.118) | 0.314 |
| Highest | 1.73 (1.42 – 2.11) | **< 0.001** | 1.01 (0.54 – 1.88) | 0.968 |
| **Religion** |  |  |  |  |
| Islam | 1 |  | 1 |  |
| Christianity | 1.13 (0.96. – 1.34) | 0.147 | 0.73 (0.55 – 0.97) | **0.032** |
| **Had COVID-19 test done before** |  |  |  |  |
| No | 1 |  | 1 |  |
| Yes | 4.95 (3.54 – 6.92) | < **0.001** | 4.32 (3.13 – 5.98) | < **0.001** |
| **Know someone previously diagnosed with COVID-19** |  |  |  |  |
| No | 1 |  | 1 |  |
| Yes | 2.25 (1.76 – 2.87) | < **0.001** | 1.41 (1.06 -1.87) | **0.020** |

<sup>a</sup>Adjusted for state clustering

**Table 4:** Factors associated with COVID-19 vaccine uptake among healthcare workers.

| **Characteristics** | **Crude OR (95% CI)** | **p-value** | **Adjusted OR (95% CI)** | **p-value** |
| --- | --- | --- | --- | --- |
| **Age** | 1.04 (0.98 – 1.11) | 0.153 | 1.06 (1.00 – 1.13) | **0.046** |
| **Sex** |  |  |  |  |
| Female | 1 |  | 1 |  |
| Male | 2.87 (0.37 – 22.4) | 0.314 | 0.93 (0.10 – 8.94) | 0.948 |
| **Religion** |  |  |  |  |
| Islam | 1 |  | 1 |  |
| Christianity | 0.41 (0.11 – 1.51) | 0.180 | 0.59 (0.15 – 2.33) | 0.453 |
| **Had COVID-19 test done before** |  |  |  |  |
| No | 1 |  | 1 |  |
| Yes | 6.23 (0.80 – 48.50) | 0.080 | 8.25 (1.04 – 65.28) | **0.046** |
| **Know someone previously** |  |  |  |  |
| **diagnosed with**<br>**COVID-19** |  |  |  |  |
| No | 1 |  | 1 |  |
| Yes | 0.64 (0.21 – 2.01) | 0.450 | 0.40 (0.11 – 1.43) | 0.159 |
| **State** |  |  |  |  |
| Lagos | 1 |  | 1 |  |
| Oyo | 0.45 (0.14 – 1.52) | 0.200 | 0.32 (0.09 – 1.14) | 0.079 |
| Jigawa<sup>a</sup> | - | - | - | - |

<sup>a</sup>There was a perfect prediction as all were vaccinated

### Individual perceptions influencing vaccine acceptance

In both the community members and healthcare workers, diverse perceptions regarding COVID-19 illness and vaccination exist. Some community members questioned its existence, government involvement, and media portrayal, with 29.5% believing COVID-19 was exaggerated by the media and 27.8% of community members believing that Africans were immune to the virus. On the other hand, 34.2% of respondents considered COVID-19 to be a death sentence. Healthcare workers also held differing views, with 22.0% believing in African immunity and 17.7% considering COVID-19 to be a death sentence (Table 5).

**Table 5:** Respondents’ perception about COVID-19 (N = 2602)

| **Characteristics** | **Total** | **Community members (N=2206)** | **Healthcare workers (N=396)** |
| --- | --- | --- | --- |
| **COVID-19 is not real** |  |  |  |
| Agree | 526 (20.2) | 467 (21.2) | 59 (14.9) |
| Disagree | 1828 (70.3) | 1496 (67.8) | 332 (83.8) |
| Don't know | 248 (9.5) | 243 (11.0) | 5 (1.3) |
| **There is no COVID-19 in Nigeria** |  |  |  |
| Agree | 367 (14.1) | 336 (15.2) | 31 (7.8) |
| Disagree | 1925 (74.0) | 1568 (71.1) | 357 (90.2) |
| Don't know | 310 (11.9) | 302 (13.7) | 8 (2.0) |
| **COVID-19 is a scam by the government** |  |  |  |
| Agree | 334 (12.8) | 312 (14.1) | 22 (5.6) |
| Disagree | 1698 (65.3) | 1361 (61.7) | 337 (85.1) |
| Don't know | 570 (21.9) | 533 (24.2) | 37 (9.3) |
| **COVID-19 is exaggerated by the media** |  |  |  |
| Agree | 714 (27.4) | 651 (29.5) | 63 (15.9) |
| Disagree | 1335 (51.3) | 1044 (47.3) | 291 (73.5) |
| Don't know | 553 (21.3) | 511 (23.2) | 42 (10.6) |
| **COVID-19 is God's punishment to mankind** |  |  |  |
| Agree | 806 (31.0) | 730 (33.1) | 76 (19.2) |
| Disagree | 1194 (45.9) | 941 (42.7) | 253 (63.9) |
| Don't know | 602 (23.1) | 535 (24.2) | 67 (16.9) |
| **COVID-19 is a biological weapon** |  |  |  |
| Agree | 470 (18.1) | 418 (18.9) | 52 (13.1) |
| Disagree | 1343 (51.6) | 1063 (48.2) | 280 (70.7) |
| Don't know | 789 (30.3) | 725 (32.9) | 64 (16.2) |
| **The virus was designed by the pharmaceutical industry to sell drugs** |  |  |  |
| Agree | 293 (11.3) | 272 (12.3) | 21 (5.3) |
| Disagree | 1523 (58.5) | 1207 (54.7) | 316 (79.8) |
| Don't know | 786 (30.2) | 727 (33.0) | 59 (14.9) |
| **Africans are immune to COVID-19** |  |  |  |
| Agree | 699 (26.9) | 612 (27.8) | 87 (22.0) |
| Disagree | 1462 (56.2) | 1174 (53.2) | 288 (72.7) |
| Don't know | 441 (16.9) | 420 (19.0) | 21 (5.3) |
| **COVID-19 is disease of the elites** |  |  |  |
| Agree | 488 (18.8) | 448 (20.3) | 40 (10.1) |
| Disagree | 1785 (68.6) | 1437 (65.1) | 348 (87.9) |
| Don't know | 329 (12.6) | 321 (14.6) | 8 (2.0) |
| **COVID-19 can affect both young and old** |  |  |  |
| Agree | 2150 (82.6) | 1793 (81.3) | 357 (90.1) |
| Disagree | 242 (9.3) | 210 (9.5) | 32 (8.1) |
| Don't know | 210 (8.1) | 203 (9.2) | 7 (1.8) |
| **COVID-19 is death sentence** |  |  |  |
| Agree | 825 (31.7) | 755 (34.2) | 70 (17.7) |
| Disagree | 1412 (54.3) | 1102 (50.0) | 310 (78.3) |
| Don't know | 365 (14.0) | 349 (15.8) | 16 (4.0) |

The primary reason for COVID-19 vaccine uptake cited by most community members (88.9%) and healthcare workers (96.1%) was to protect themselves against COVID-19 infection. Compliance with government directives was mentioned by 36.5% of community members and 38.1% of healthcare workers. Additionally, a portion of the respondents, particularly healthcare workers (36.7%), reported taking the vaccine to encourage others to do the same, whereas a smaller percentage (1.3%) indicated doing so because others were taking it (due to social influence). Some community members (15.3%) received the vaccine for travel purposes (Table 6).

**Table 6:** Reasons for COVID-19 vaccine acceptance among respondents that received any dose of vaccination.

| **Reasons for taking vaccine<sup>a</sup>** | **Community members/patients**<br>**(n = 957)** | **Healthcare workers**<br>**(n = 383)** |
| --- | --- | --- |
| To protect me against COVID-19 infection | 832 (86.9) | 368 (96.1) |
| To comply with the government directive | 349 (36.5) | 146 (38.1) |
| To encourage others to take it | 117 (12.2) | 144 (37.6) |
| For travel purposes | 146 (15.3) | 66 (17.2) |
| Because others are taking it | 33 (3.5) | 5 (1.3) |
| Because it is free | 31 (3.2) | 11 (2.9) |
| Have had COVID-19 infection before | 1 (0.1) | 3 (0.8) |
| To comply with workplace/boss directive | 10 (1.0) | 1 (0.3) |
| Because I will be paid/given cash | 2 (0.2) | - |
| Others | 6 (0.6) | 2 (0.5) |

<sup>a</sup> Respondents could provide multiple responses

While the quantitative data primarily highlighted self-protection and compliance with directives, qualitative insights delve deeper into individual factors. Participants expressed reasons such as setting examples for others, aligning with influential figures, ensuring job security, and protecting themselves and their families. For instance, one healthcare worker expressed,

> *“I saw our President; he took his own. Our state first man, he took his own. Our board, and my oga here took her own. So, who am I”* (HCW 002 Oyo)

Some community members noted,

> *“Why I went was that government workers won’t be able to work without it”* (Nursing mother 002 Oyo)

### Hesitancy and experiences with COVID-19 vaccination

The reasons for vaccine hesitancy were diverse, including confidence-related barriers, particularly fear of side effects (32.6%), pregnancy-related factors (25.9%), and convenience-related barriers (13.5%). Non-uptake of the second dose was driven by complacency-related barriers (51.6%), convenience-related barriers (30.9%), and pregnancy-related factors (16.8%). Lack of uptake of booster doses was influenced by convenience-related barriers (32.5%), health system factors (29.8%), and information-related barriers (18.0%) (Fig 1 and S3 Table).

**Fig 1.** ^a^ Reasons for COVID-19 vaccine hesitancy among unvaccinated eligible respondents. (Note: “Other reasons” was indicated as barriers by 1.1% of the respondents for no dose, 0.8% for no second dose, and 0.3% for no booster dose. Also, conspiracy theory/rumour/misconception was cited as barrier by 1.1% of the respondents that didn’t receive any dose of the vaccine. ^a^ Respondents could provide multiple responses)

The qualitative analysis revealed additional insights into respondents’ COVID-19 vaccine hesitancy, with participants expressing concerns about vaccine safety, citing misinformation and mistrust in the government.

> *“Lack of trust in the government, they think they want to reduce the number of people in the world and Nigeria, some say it is for family planning”* (Caregiver 001 Jigawa)

> *“There is wrong information on COVID 19 vaccine, wrong news of side effects and so on. Some people believed that government is using the COVID-19 vaccine to steal more money for their families”* (HCW 03 Oyo)

Some mentioned fear of adverse reactions or doubts regarding the vaccine’s effectiveness.

> *"It has reaction, some will be sleepy all day and night, some fever, some dizziness”*

(Caregiver 003 Jigawa)

> *“There was a community we went for vaccination, and they said, a man died after being vaccinated and the community ever since then refused to receive vaccine”* (HCW 04 Jigawa)

Others highlighted logistical challenges, such as the inconvenience of vaccine administration or difficulties accessing healthcare facilities.

> *“I took an excuse from my workplace that I want to go and take covid vaccine, right? On getting there, I was told I’ll wait for 3/4 hours. Do you think I will wait? I won’t like to risk my job”* (Community member 002 Oyo)

The need for spousal approval and the belief that childhood vaccinations are sufficient for protection against diseases were also identified from the qualitative data.

> *“Some of us will have to take permission from our husbands”* (Nursing mother 001 Jigawa)

> *“I don’t like it; I believe that all the vaccines that I have taken in my childhood is enough to sustain me”* (Nursing mother 03 OYO)

### Trust in vaccines and future intentions regarding vaccination

Despite COVID-19 vaccine hesitancy, particularly among community members, a significant proportion of respondents expressed overall trust in vaccine safety and efficacy, indicating a willingness to vaccinate against other diseases. There was little difference between unvaccinated and vaccinated respondents in their belief in vaccine safety (94.9% vs. 98.2%) and efficacy (95.5% vs. 98.2%) of routine vaccines. For COVID-19 vaccine recipients, their belief in vaccine safety and efficacy increased with COVID-19 to 99.2% and 99.0%, respectively.

Furthermore, the majority of vaccine recipients (99.3%) expressed willingness to vaccinate their children against childhood illnesses, 97.8% were willing to take a vaccine against diseases like hepatitis, 97.0% were willing to take a vaccine against human papillomavirus (HPV), and 98.7% were willing to take vaccines in the future in case of another similar outbreak (S4 Table).

## Discussion

In this study, we examined COVID-19 vaccination patterns and factors influencing vaccine uptake and perceptions of the COVID-19 pandemic among community members and healthcare workers in Nigeria. We found significant disparities in vaccination rates and underscored the multifaceted nature of vaccine acceptance and hesitancy. While HCWs exhibited high initial vaccine uptake, the drop-off was high, with only one-third completing the booster dose. In contrast, over half of the community members remain unvaccinated, highlighting ongoing challenges in achieving comprehensive COVID-19 vaccination coverage, particularly in the WHO African region, which has failed to meet the 70% vaccination target.(10) These findings are consistent with previous reports of suboptimal COVID-19 vaccine uptake in Nigeria.(29–31)

We found several socio-demographic factors positively associated with vaccine acceptance among community members, including older age, male sex, a history of COVID-19 testing, and knowing someone previously diagnosed with COVID-19. These findings align with the understanding that older individuals at an increased risk of severe COVID-19 outcomes are more likely to seek vaccination.(32) Knowing someone affected by COVID-19 served as a motivational factor for vaccination, likely due to heightened awareness of the disease’s severity.(33) For HCWs, older age and previous COVID-19 testing were associated with vaccine uptake, suggesting that direct exposure to COVID-19, whether through professional experience or personal testing, significantly influences vaccine acceptance among HCWs.(33) The heightened vulnerability of older HCWs likely drives their proactive health behaviours, including vaccination.

Community members who practiced Christianity had lower vaccination uptake. This finding resonates with a qualitative study among faith leaders in Addis Ababa, Ethiopia.(34) Individual perceptions also play a significant role in shaping vaccine acceptance. Both community members and HCWs expressed scepticism about COVID-19’s existence, notions of African immunity, and misconceptions about the disease being exaggerated by the media. Prior studies have shown that public perceptions and beliefs significantly influence compliance with preventive measures and contribute to vaccine hesitancy.(35–37) A study conducted by the Africa CDC on COVID-19 vaccine perception across 15 countries, including Nigeria, found misconceptions, mistrust, misinformation, and fears about vaccine safety.(38) Rumours and myths surrounding vaccine development by developed nations as a population reduction strategy in Africa, with claims that these vaccines cause infertility, preceded COVID-19 and have led to mistrust among the public.(30,34,39–41) These underscore the need for comprehensive strategies to rectify misbeliefs and misinformation and the importance of targeted engagement with community heads and religious leaders who are highly regarded in African communities and among their congregations to address misconceptions and enhance vaccine confidence.

COVID-19 vaccination programs were a cornerstone of the pandemic response, both to curb outbreaks and long-term instruments to prevent subsequent waves of the COVID-19 pandemic.(42) Key obstacles to vaccination reported in our quantitative findings included confidence-related issues, especially fear of side effects,(30,40,41) pregnancy-related concerns, and convenience-related barriers. The qualitative data provided additional barriers, such as inconveniences related to vaccine administration, the need for spousal approval, and belief in the sufficiency of childhood vaccinations. Confidence barriers tended to diminish with subsequent vaccine doses, suggesting a gradual increase in trust over time. However, respondents reported higher levels of complacency, convenience-related issues, health system factors, and information barriers for subsequent doses than for the initial dose. Complacency was particularly high for the second dose, indicating a need for continuous awareness campaigns. Convenience-related barriers, such as prolonged waiting times and limited access, were significant deterrents, consistent with the findings of Agha (2021), who reported only 32% of respondents found it easy to obtain COVID-19 vaccination for themselves,(43) reinforcing the need for streamlined vaccination processes.

Despite these barriers, we found several motivating factors, including protection against infection, compliance with government directives, encouragement from others, and travel purposes. We also saw the influence of social factors, such as seeing influential figures like the president and local leaders take vaccine-motivated others to follow suit. Among HCWs, taking the vaccine to set an example for others and fulfil job requirements was particularly evident. The findings of this study align with those of a qualitative study conducted in Ethiopia, where religious leaders who are highly respected in their communities and congregations not only promoted preventive measures during the pandemic but also received vaccinations publicly to build trust among the people.(34)

## Limitations

We had three key limitations in our study. Firstly, the reliance on self-reported data may have introduced bias, as participants might have over-reported and under-reported their vaccination status, perceptions and experiences due to social desirability or recall bias. Secondly, data collection was limited to community members and healthcare workers at primary healthcare facilities, meaning the perspectives and experiences of other community members without facility engagement during the data collection period were not explored. Finally, logistical constraints and variations in vaccine availability during the study period could have influenced vaccination patterns, thus affecting the results. Despite these limitations, utilizing a mixed-method approach in our study provided a unique opportunity to comprehend the intricacies of COVID-19 immunization programs in Nigeria and glean insights applicable during another pandemic.

## Conclusion

Our study highlights preventable obstacles impacting vaccination intentions, emphasizing the need for the Nigerian Government to enhance its implementation program. This involves bolstering vaccine literacy through effective public communication strategies and ensuring consistent accessibility of vaccines when needed. The findings underscore the importance of addressing religious beliefs and socio-demographic factors significantly influencing vaccine uptake. Additionally, overcoming barriers such as misinformation, mistrust in the government, and logistical challenges is crucial. Involving influential figures and trusted individuals in shaping messages and information sharing could be proactive measures to instill confidence and ensure reliable information spreads. These lessons are important to prepare for the next pandemic.

## Data Availability

Data cannot be shared publicly because they contain personal information of the participants. Data are however available from the researcers who meet criteria for access to confidential data

## Acknowledgement

We thank the data collectors, healthcare workers and community members for their time and support.

