## Supplemental materials for "Exploring COVID-19 Pandemic Perceptions and Vaccine Uptake among Community Members and Primary Healthcare Workers in Nigeria: A Mixed Methods Study"

**S1 Figure:** **Participants inclusion in data collection at facilities**

**Approached**

(n = 2734)

**No consent**

(n = 5)

**Consent**

(n = 2729)

**Not eligible** (n = 127)

- **HCWs** not involved in Immunization (30)
- **Community members** that recently relocated or not primary residence/required urgent medical attention (97)
- Not primary residence (

**Eligible**

(n = 2602)

**Community members**

(n = 2206)

**Healthcare workers**

(n = 396)

**S2 Table: Qualitative - Socio-demographic characteristics of respondents**

| Participants characteristics | Community Members | | Healthcare providers | |
| --- | --- | --- | --- | --- |
|  | Oyo (n=8) | Jigawa (n=8) | Oyo (n=6) | Jigawa (n=8) |
| Sex |  |  |  |  |
| Male | 2 | 1 | 0 | 6 |
| Female | 6 | 7 | 6 | 2 |
| Religion |  |  |  |  |
| Islam | 5 | 8 | 3 | 8 |
| Christianity | 3 | 0 | 3 | 0 |
| Level of education |  |  |  |  |
| Primary/no formal | 0 | 4 | 0 | 0 |
| Secondary | 2 | 3 | 1 | 0 |
| Tertiary | 6 | 0 | 5 | 8 |

**S3 Table:** **Reasons for COVID-19 vaccine hesitancy among unvaccinated eligible respondents**

| **Reasons for not taking vaccine^a^** | **No dose^b^** | | **No second dose^b^** | | **No booster dose^b^** | |
| --- | --- | --- | --- | --- | --- | --- |
|  | **Community members (n = 1234)** | **Healthcare workers (n =13)** | **Community members (n =233)** | **Healthcare workers (n =23)** | **Community members (n=174)** | **Healthcare workers (n =121)** |
| **Confidence related barriers** |  |  |  |  |  |  |
| Fear of side effects/side effects from previous dose | 302 (24.5) | 4 (30.8) | 33 (14.2) | 2 (8.7) | 4 (2.3) | 3 (2.5) |
| Lack of trust in the quality of the vaccine | 98 (7.9) | 1 (7.7) | - | - | 1 (0.6) | 1 (0.8) |
| Lack of trust in the government | 67 (5.4) | - | - | - | - | - |
| **Conspiracy theory, rumour, and misconception** |  |  |  |  |  |  |
| It is a mark of the beast | 7 (0.6) | - | - | - | - | - |
| It will lead to infertility | 7 (0.6) | - | - | - | - | - |
| **Complacency related barriers** |  |  |  |  |  |  |
| Belief there is no COVID-19 | 31 (2.5) | - |  |  |  |  |
| I don’t need the vaccine/previous dose(s) taken is sufficient | 117 (9.5) | 2 (15.4) | 114 (48.9) | 15 (65.2) | 14 (8.1) | 22 (18.2) |
| Loss of vaccine card |  |  | 8 (3.4) | - | 5 (2.9) | - |
| **Convenience related barriers** |  |  |  |  |  |  |
| Long waiting time/busy | 77 (6.2) | 4 (30.8) | 15 (6.4) | 4 (17.4) | 18 (10.3) | 25 (20.7) |
| Travel out of town/I need to go to where to take second or booster dose | 22 (1.8) | - | 34 (14.6) | 7 (30.4) | 33 (19.0) | 13 (10.7) |
| Due to sickness | 19 (1.5) | - | 3 (1.3) | - | 2 (1.2) | 3 (2.5) |
| Lack of access | 45 (3.7) | - | 16 (6.9) | - | 2 (1.2) | 1 (0.8) |
| **Information related barriers** |  |  |  |  |  |  |
| Not aware of the COVID-19 vaccine/second or booster dose | 88 (7.1) | - | 13 (5.6) | - | 47 (27.0) | 6 (5.0) |
| **Pregnancy related factors** |  |  |  |  |  |  |
| Due to pregnancy | 245 (19.9) | - | 39 (16.7) | 1 (4.4) | 6 (3.5) | 2 (1.7) |
| Due to breastfeeding | 108 (8.8) | 1 (7.7) | 3 (1.3) | - |  |  |
| **Socio-cultural and political related factors** |  |  |  |  |  |  |
| It is against my religious belief | 37 (3.0) | - | - | - | - | - |
| Husband did not give permission | 9 (0.73) | - | - | - | - | - |
| Not a beneficiary of political interventions (Cash Transfer policy, government empowerment programme) | 10 (0.8) |  | 1 (0.4) | - | - | - |
| Gender discrimination | 1 (0.1) | - | - | - | - | - |
| **Health systems factor** |  |  |  |  |  |  |
| Vaccine is not available | 10 (0.8) | 1 (7.7) | 30 (12.9) | 2 (8.7) | 45 (25.9) | 43 (35.5) |
| **Others** | 14 (1.1) | **-** | 2 (0.9) | - | - | 1 (0.8) |

**^a^ Respondents could provide multiple responses**

**^b^ Excluding respondents that are not eligible for dose, dose not required or did not indicate reasons**

**S4 Table: Respondents’ perception about COVID-19 vaccine, recommendation & willingness to take other vaccines.**

| **Characteristics** | **Total** | **Community members/patients (N= 1769)** | **Healthcare workers (N=396)** |
| --- | --- | --- | --- |
|  | **N (%)** | **N (%)** | **N (%)** |
| **If NO to receiving any dose COVID-19 vaccine (N = 1247)** | | | |
| **Do you believe in vaccine safety?** |  |  |  |
| Yes | 1183 (94.9) | 1171 (94.9) | 12 (92.3) |
| No | 64 (5.1) | 63 (5.1) | 1 (7.7) |
| **Do you believe in vaccine efficacy?** |  |  |  |
| Yes | 1191 (95.5) | 1179 (95.5) | 12 (92.3) |
| No | 56 (4.5) | 55 (4.5) | 1 (7.7) |
| **If YES to receiving any dose COVID-19 vaccine (N = 1349)** | | | |
| **Before the covid-19 pandemic, did you believe in vaccine safety?** |  |  |  |
| Yes | 1324 (98.2) | 945 (97.8) | 379 (99.0) |
| No | 25 (1.8) | 21 (2.2) | 4 (1.0) |
| **Before the covid-19 pandemic, did you believe in vaccine efficacy?** |  |  |  |
| Yes | 1326 (98.3) | 949 (98.2) | 377 (98.4) |
| No | 23 (1.7) | 17 (1.8) | 6 (1.6) |
| **Based on your experience with the covid-19 vaccination, do you believe in vaccine safety?** |  |  |  |
| Yes | 1338 (99.2) | 957 (99.1) | 381 (99.5) |
| No | 11 (0.8) | 9 (0.9) | 2 (0.5) |
| **Based on your experience with the covid-19 vaccination, do you believe in vaccine efficacy?** |  |  |  |
| Yes | 1335 (99.0) | 954 (98.8) | 381 (99.5) |
| No | 14 (1.0) | 12 (1.2) | 2 (0.5) |
| **Based on your experience with the covid-19 vaccination will you recommend covid-19 vaccine to someone else?** |  |  |  |
| Yes | 1330 (98.6) | 949 (98.2) | 381 (99.5) |
| No | 19 (1.4) | 17 (1.8) | 2 (0.5) |
| **Based on your experience with the covid-19 vaccination will you allow your child to take routine available vaccines against common childhood illnesses?** |  |  |  |
| Yes | 1340 (99.33) | 960 (99.4) | 380 (99.2) |
| No | 9 (0.67) | 6 (0.6) | 3 (0.8) |
| **Based on your experience with the covid-19 vaccination are you willing to take vaccine against disease like hepatitis infection?** |  |  |  |
| Yes | 1316 (97.8) | 938 (97.4) | 378 (98.7) |
| No | 30 (2.2) | 25 (2.6) | 5 (1.3) |
| **Based on your experience with the covid-19 vaccination are you willing to take vaccine against human papilloma virus?** |  |  |  |
| Yes | 1309 (97.0) | 932 (96.5) | 377 (98.4) |
| No | 40 (3.0) | 34 (3.5) | 6 (1.6) |
| **Based on your experience with the covid-19 vaccination are you willing to take a vaccine in the future in case of outbreak similar to covid-19 pandemic?** |  |  |  |
| Yes | 1332 (98.7) | 951 (98.4) | 381 (99.5) |
| No | 17 (1.3) | 15 (1.6) | 2 (0.5) |

**S5: Healthcare provider In-depth Interview guide**

We will be conducting interviews with healthcare providers who work in primary health facilities that offer routine immunization and COVID-19 immunization services

1. Clinical context

- Can you tell me about a typical immunization clinic (routine immunization) in your setting?
  - Probe: What sort of activities are done? How many children do you see on average?
  - Probe frequency of clinics; clinic duration
  - Probe preparation done prior to the clinic
- If the facility provides COVID-19 immunization services:
  - Probe: What sort of activities are done? What role does the participant perform? How many recipients do you attend to on average per day?
  - Probe frequency of clinics, clinic hours
  - Probe preparation done prior to the clinic (COVID-19 vaccination)
  - If not, probe why it does not offer COVID-19 immunization services
  - What are the processes involved to receive COVID-19 vaccine
  - What is working well with COVID-19 immunization delivery in Nigeria
  - What is not working well with COVID-19 immunization delivery in Nigeria

2. Perception of integrated COVID-19/routine immunization deliveries
   a. What is your opinion of providing COVID-19 immunization with other routine immunization services in your facility as recommended by the NPHCDA?
   b. Does it increase coverage for COVID-19 vaccine? How?
   c. How do healthcare providers consider that approach? Does it help your work? How?
   d. NPHCDA also recommends screening for hypertension and diabetes alongside COVID-19 vaccination. To what extent this is implemented at your facility? How is this approached perceived by the community members.
3. Existing challenges with routine immunization services
   a. What is working well with routine immunization delivery in Nigeria
   b. What is not working well with routine immunization delivery in Nigeria
4. Impact of COVID-19 immunization services on routine immunization service
   a. In what ways has COVID-19 immunization programme affected routine immunization delivery in your facility?
      i. Probe positive impacts if any
      ii. Probe negative impact if any
      iii. Has COVID-19 affected people’s perception of vaccines generally in Nigeria? How
   b. What effects does routine immunization have on COVID-19 immunization programme?
      i. Tell me more, can you give examples?
5. COVID-19 vaccine recipients
   a. Have you received at least a dose of the COVID-19 vaccine before? If yes share your experience (registration, verification, and actual inoculation)
      i. Probe motivation for acceptability or hesitancy
         1. Confidence in vaccine
         2. Complacency
         3. Convenience
      ii. Comment on the steps involved to get vaccinated. Were the steps necessary? Why?
      iii. Experiences post-vaccination, side effects if any? How were they managed?
      iv. Were the side effects expected? How?
      v. Perceived difference between COVID-19 immunization services and RI
      vi. Challenges encountered
      vii. Why do some people receive the COVID-19 vaccine?
      viii. Why are other people not willing to accept the vaccine?
6. Do you have any other things to tell me?
7. Thank you for your time

**S6: Community members In-depth Interview guide**

We will be conducting interviews with non-healthcare providers who present in primary health facilities

1. Getting to know the participants

- Tell me about yourself: where you live, work you do, marital status, religion and tribe

2. Tell me your perception of immunization generally? Why
   a. What vaccine(s) have you received in your adulthood? Why?
   b. Do you take your children for immunization? Why
   c. Many under-five children are not fully immunized in Nigeria. What could be responsible for this?
   d. Any other challenges regarding immunization programme for children in Nigeria?
   e. What is working well with the ways routine immunization for children is delivered in Nigeria?
   f. What is not working well with the children’s routine immunization?
3. COVID-19 vaccination experience
   a. Have you received at least a dose of the COVID-19 vaccine before? If yes, when and how many doses have you received? Share your experience (registration, verification, and actual inoculation), and how satisfied were you with the entire process?
      i. Probe motivation for acceptability or hesitancy
      ii. Why do some people receive the COVID-19 vaccine?
      iii. Why are other people not willing to accept the vaccine?
      iv. Comment on the steps involved to get vaccinated. Were the steps necessary? Why?
      v. What were your experiences post-vaccination, side effects if any? How were they managed? Were the side effects expected? How?
      vi. What is not working well with COVID-19 immunization delivery in Nigeria
      vii. What do you consider to be working well with COVID-19 immunization in Nigeria
4. Impact of COVID-19 immunization services on routine immunization service
   a. Of the challenges you discussed so far, which ones are peculiar to:
      i. Routine immunization? How
      ii. COVID-19 vaccine programme? How?
   b. Based on your opinion and experiences, in what ways has the COVID-19 immunization programme affected immunization for children in your setting?
      i. Probe negative impact if any
      ii. Probe positive impacts if any
      iii. Has COVID-19 immunization programme shaped how people perceive vaccines generally?
   c. We have talked about effects of COVID-19 vaccine on routine immunization. Now what effects does routine immunization have on COVID-19 immunization programme?
      i. Tell me more, can you give examples?
